# Does smoking during pregnancy influence offspring handedness? Extending gene-by-environment Mendelian randomisation to include polygenic risk scores

**DOI:** 10.1101/2024.10.14.24315490

**Authors:** Daisy CP Crick, Sarah Medland, George Davey Smith, David Evans

## Abstract

Hand preference first appears *in utero*, yet twin studies and GWAS show that the majority of variance in hand preference is explained by environmental factors. Using UK Biobank data and multivariable logistic regression to test associations between potential causes of handedness and offspring hand preference, we found maternal smoking during pregnancy increased the probability of being right-handed after adjustment for covariates. Using a proxy gene-by-environment (GxE) Mendelian randomization design we investigated the potential causal effect of maternal smoking during pregnancy on offspring handedness. We used rs16969968 in the *CHRNA5* gene and a polygenic risk score of genome-wide significant smoking-heaviness variants to proxy smoking behaviour. We stratified based on reported maternal smoking during pregnancy because, regardless of genotype, any causal effect of maternal smoking on offspring handedness should only manifest in individuals whose mothers smoked during pregnancy. The GxE MR analyses found no causal effect of maternal smoking during pregnancy on offspring hand-preference. Our study contributes to the understanding of hand preference and its potential early-life determinants. However, the main factors contributing to variation in hand preference remain unresolved.

---

The majority of people (90%) are right-handed, meaning that they use their right hand for complex manual tasks such as writing [1, 2]. The rest of the population is split into those who are left-handed (∼10%) and a small fraction of individuals who are ambidextrous and can use either hand equally well (∼1%) [2]. The first signs of hand preference and motor asymmetry in humans emerge at around 15 weeks gestation in the form of preferential lateralized thumb sucking behaviours *in utero* [3]. By 5-6 months post-partum, infants exhibit clear hand preferences for target-directed behaviours [4] which then tend to remain stable throughout childhood [5]. Certain early life influences are thought to affect hand preference. For instance, males are more likely than females to be left-handed [6], and lower birthweight and antenatal complications [7] are associated with a higher prevalence of left-handedness [8]. Cultural pressures also appear to impact hand preference [9], evident through the lower frequency of left-handers in Asia compared to North America or Europe [9] and through the steady increase in left-handedness over time as societal demands for right handedness diminish [10]. Whilst observational, associations have been reported between left-handedness/ambidexterity and a number of neurological and psychiatric disorders such as dyslexia [11], schizophrenia [12] [13], depression [14], bipolar disorder [15], and migraines [16].

There is now overwhelming evidence that handedness is at least partially genetically determined and probably highly polygenic (i.e. as opposed to being the result of a single genetic variant or a small number of genetic variants [17]). In the largest genome-wide association study (GWAS) of handedness to date, Cuellar-Partida *et al*. identified 48 genetic variants (single nucleotide polymorphisms; SNPs) associated with individual hand preference [18]. Of these, 41 influenced a person’s likelihood of being left-handed/right-handed, while the other seven were associated with using both hands for tasks. In addition, the authors estimated that additive genetic factors explained ∼12% (95% CI: 7.20, 17.70) of the variance in the underlying liability of being left-handed [18] by examining genome-wide identity-by-descent sharing and handedness concordance in close relatives in the UK Biobank [19]. Although this figure is slightly lower than estimates from the largest twin study of handedness to date (∼25% heritability (95% CI: 15.69, 29.51) [20], the evidence clearly shows that (unshared) environmental, rather than genetic, factors are responsible for most of the variation in handedness. Given that hand preference appears early in development, the corollary is that early life environmental exposures (i.e. *in utero* and shortly after birth) are likely to be important determinants of handedness.

One potential cause for a shift towards left-handedness is foetal hypoxia as a result of maternal smoking during pregnancy [21]. Maternal smoking during pregnancy can produce foetal hypoxia in several ways: it can reduce oxygen supply to the foetus due to the production and binding of carbon monoxide (a constituent of tobacco smoke) to haemoglobin [22]; nicotine can reduce blood flow to the foetus due to vasoconstriction [22], and/or increased blood viscosity (i.e. as a result of red blood cells increasing in size to compensate for reduced oxygen) [23]; and smoking can damage the placenta which can then reduce blood flow and therefore the amount of oxygen and nutrients delivered to the developing foetus [24]. Thus, because left-handedness is associated with gestational and birth complications [7] and many birth complications are a result of prenatal or perinatal hypoxia [21], foetal hypoxia due to maternal smoking may also increase the frequency of left-handedness.

Studies investigating the relationship between maternal smoking and offspring handedness are sparse and conflicting [21] [25]. In addition, observational studies are susceptible to bias and confounding and any reported associations may not reflect causal relationships between the phenotypes [26]. Mendelian randomization (MR) studies [27], in which genetic variants are used as instrumental variables (IVs) to proxy the exposure of interest, provide an alternative way to investigate potential causal relationships between traits [27, 28]. MR studies are, in theory, less prone to confounding compared to traditional observational epidemiological studies because genetic variants segregate randomly and assort independently of potential genetic and environmental confounders [28].

In the present study we used data from the UK Biobank (UKB) to investigate a possible causal effect of maternal smoking during pregnancy on offspring handedness. We first examined the association between maternal smoking and other early life factors on offspring hand preference using multivariable logistic regression. We then used a proxy gene by environment (GxE) Mendelian randomization method [29] where an individual’s own genotype for smoking heaviness (i.e. at the rs16966968 variant in the *CHRNA5* gene) [30] was used to proxy the genotype of their mothers at the same locus, and therefore their mother’s smoking behaviour, to examine causality (Figure 1). If the heaviness of maternal smoking increases the likelihood of left-handed offspring, then the association between rs16966968 and handedness should only be observed in individuals who reported that their mothers smoked during pregnancy. In contrast, the presence of an association between the variant and handedness in individuals whose mothers did not smoke during pregnancy would suggest the association is a result of confounding through genetic pleiotropy [31].

**Figure 1:**
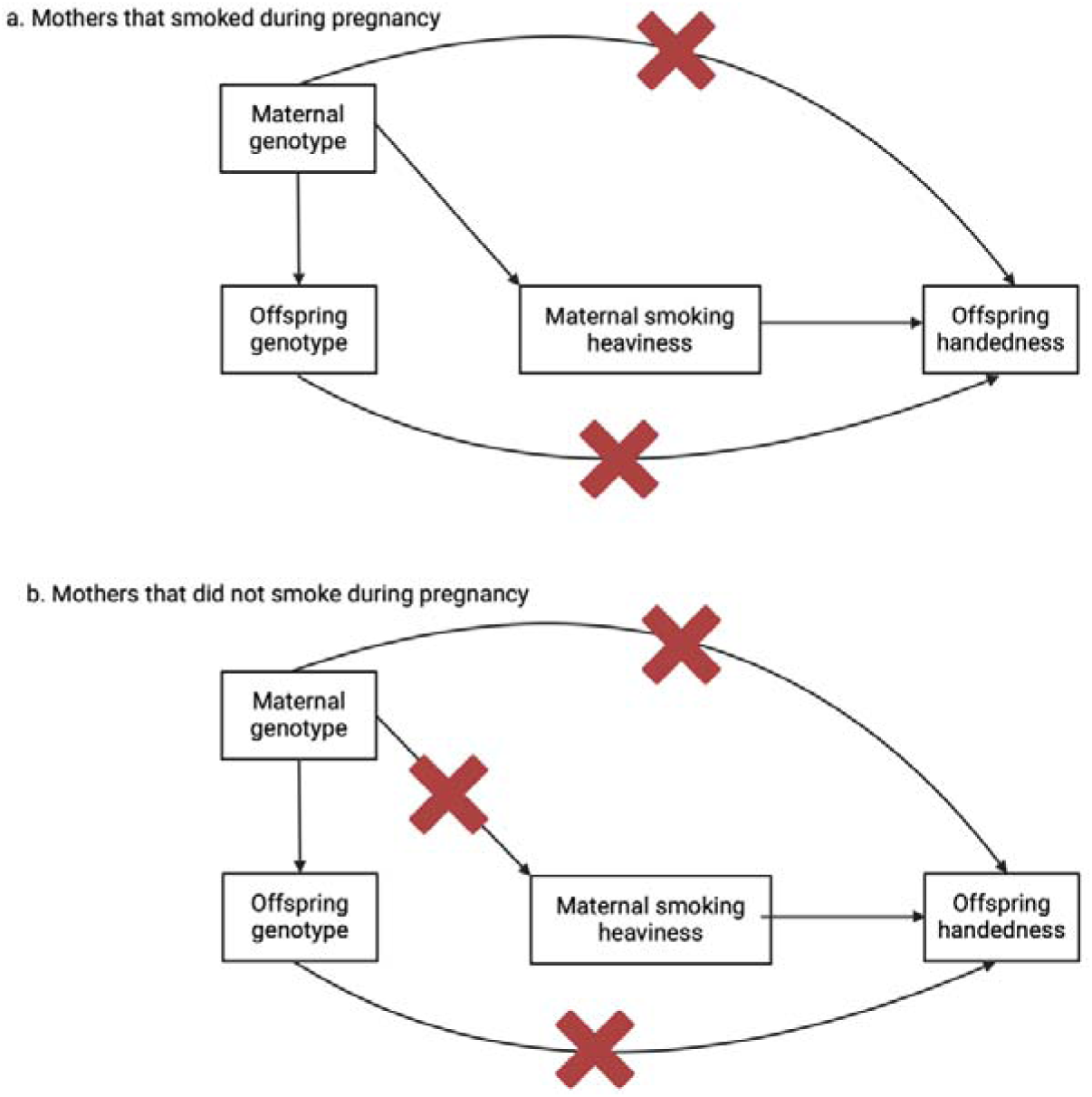
Proxy gene-by-environment Mendelian randomization conceptual framework. Offspring genotype is used as a proxy for maternal genotype and analyses are stratified on maternal smoking behaviour during pregnancy. Red crosses indicate closed pathways, for example, in figure 1b., maternal genotype for smoking heaviness did not influence smoking heaviness behaviour because the mothers did not smoke during pregnancy. Hence a red cross over the path from maternal genotype to maternal smoking heaviness. If heaviness of smoking in mothers is causal for offspring handedness, then an association between offspring genotype and offspring handedness should only be present in the children of mothers who smoked during pregnancy. The design assumes that offspring genotype only potentially associates with offspring handedness through maternal genotype and not through offspring phenotype. Since smoking initiation occurs well after hand preference manifests, offspring smoking behaviour should have no influence on offspring handedness (although this does not rule out the possibility of other pleiotropic actions on offspring handedness through the offspring genome). The design also assumes the absence of pleiotropic paths between maternal genotype and offspring handedness outside of maternal smoking behaviour. However, stratification on maternal smoking behaviour during pregnancy provides a test for the presence of horizontal pleiotropy in the maternal and/or offspring genomes. Specifically, if the above assumptions hold, the estimated causal effect of maternal smoking on offspring handedness in the offspring of mothers who did not smoke during pregnancy should be zero. If this is not the case, then the true causal effect of maternal smoking on offspring handedness (adjusted for pleiotropy) can be estimated by subtracting the causal estimate obtained in the offspring of non-smoking mothers from the causal estimate obtained in the offspring of smoking mothers. We also assume no paths from paternal genotype at the same (or correlated loci) to offspring handedness (not shown) and that the SNPs used are not associated with smoking initiation and/or termination (see the Discussion for further explication of these points).

Whilst the genotype by proxy GxE MR design provides a degree of robustness to horizontal pleiotropy, it suffers from low statistical power due to the reliance on offspring SNPs to proxy maternal genotype and because of the stratification into groups (smoking and non-smoking). We therefore extended the basic proxy GxE method to incorporate a polygenic risk score comprised of genome-wide significant SNPs for smoking heaviness (P<5×10^-8^) (PRS) [32] [33]. We reason that by extending the classic single SNP GxE design, the score may explain more of the phenotypic variance in maternal smoking heaviness, and therefore increase the power of our analysis. Additionally, the existence of a “no-relevance” control group (i.e., individuals who reported that their mothers did not smoke in pregnancy) should enable causal effect estimates to be corrected for the effect of horizontal pleiotropy.

## Results

### Early life exposures of handedness

The distribution of handedness in the whole cohort for the logistic regression analysis and the genetic subset is presented in Table 1. The distribution of characteristics in the whole cohort for the logistic regression analysis and the genetic subset is presented in Table 2. The distributions of characteristics in the retained cohort after excluding participants who have any missing values is presented in Supplementary Table 3. The pairwise relationships between predictor variables are presented in Supplementary Table 4.

**Table 1:** Distribution of responses to question about hand preference.

| **Logistic regression analysis** |  |  |  |
| --- | --- | --- | --- |
| Hand use | Males (%) | Females (%) | Total (%) |
| Right-handed | 175,965 (89.09) | 215,757 (91.19) | 391,722 (90.23) |
| Left-handed | 21,546 (10.91) | 20,836 (8.81) | 42,382 (9.76) |
| Total | 197,511 | 236,593 | 434,104 |
| **Genetic subset** |  |  |  |
| Hand use | Males (%) | Females (%) | Total (%) |
| Right-handed | 142,134 (89.02) | 170,665 (91.16) | 312799 (90.18) |
| Left-handed | 17,523 (10.98) | 16,549 (8.84) | 34072 (9.82) |
| Total | 159,657 | 187,214 | 346,871 |

**Table 2:**
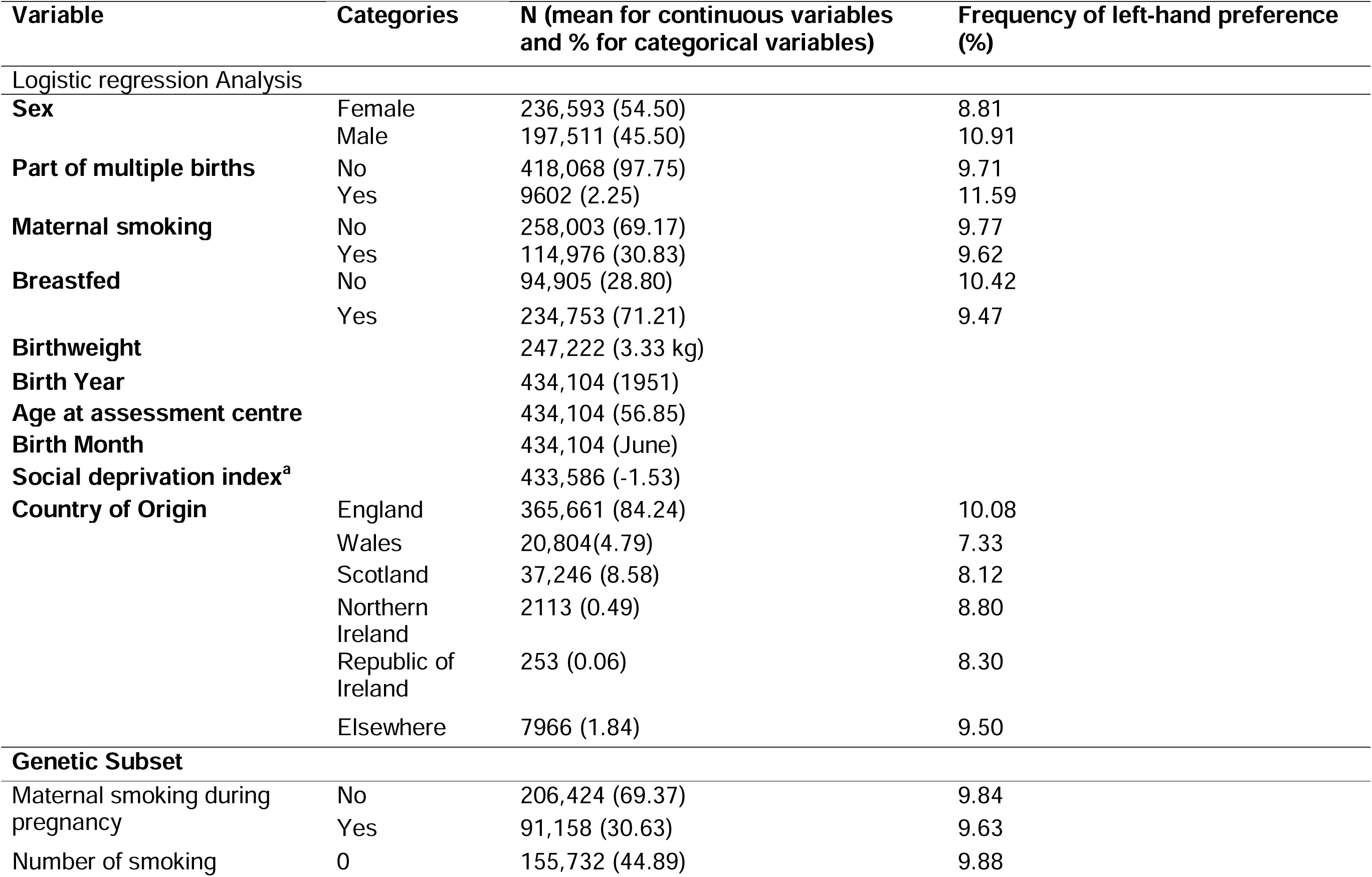
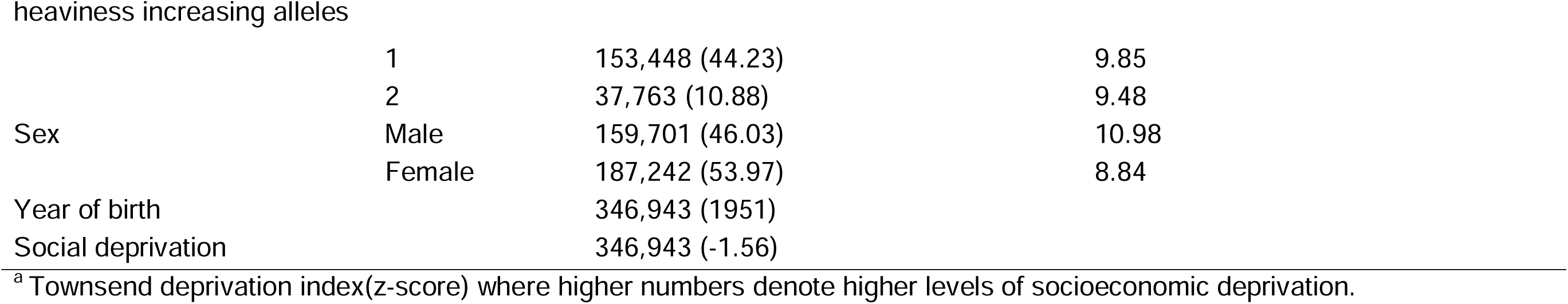
Distribution of observed characteristics at baseline assessment in UK Biobank.

| Variable | Categories | N (mean for continuous variables and % for categorical variables) | Frequency of left-hand preference (%) |
| --- | --- | --- | --- |
| Logistic regression Analysis |  |  |  |
| **Sex** | Female | 236,593 (54.50) | 8.81 |
|  | Male | 197,511 (45.50) | 10.91 |
| **Part of multiple births** | No | 418,068 (97.75) | 9.71 |
|  | Yes | 9602 (2.25) | 11.59 |
| **Maternal smoking** | No | 258,003 (69.17) | 9.77 |
|  | Yes | 114,976 (30.83) | 9.62 |
| **Breastfed** | No | 94,905 (28.80) | 10.42 |
|  | Yes | 234,753 (71.21) | 9.47 |
| **Birthweight** |  | 247,222 (3.33 kg) |  |
| **Birth Year** |  | 434,104 (1951) |  |
| **Age at assessment centre** |  | 434,104 (56.85) |  |
| **Birth Month** |  | 434,104 (June) |  |
| **Social deprivation index<sup>a</sup>** |  | 433,586 (-1.53) |  |
| **Country of Origin** | England | 365,661 (84.24) | 10.08 |
|  | Wales | 20,804(4.79) | 7.33 |
|  | Scotland | 37,246 (8.58) | 8.12 |
|  | Northern Ireland | 2113 (0.49) | 8.80 |
|  | Republic of Ireland | 253 (0.06) | 8.30 |
|  | Elsewhere | 7966 (1.84) | 9.50 |
| **Genetic Subset** |  |  |  |
| Maternal smoking during pregnancy | No | 206,424 (69.37) | 9.84 |
|  | Yes | 91,158 (30.63) | 9.63 |
| Number of smoking | 0 | 155,732 (44.89) | 9.88 |
|  | 1 | 153,448 (44.23) | 9.85 |
|  | 2 | 37,763 (10.88) | 9.48 |
| Sex | Male | 159,701 (46.03) | 10.98 |
|  | Female | 187,242 (53.97) | 8.84 |
| Year of birth |  | 346,943 (1951) |  |
| Social deprivation |  | 346,943 (-1.56) |  |

<sup>a</sup> Townsend deprivation index(z-score) where higher numbers denote higher levels of socioeconomic deprivation.

### Traditional Observational Epidemiological Analyses

Using data from UKB [19] we performed a univariable logistic regression analysis and found that there was evidence that a later year of birth, being male, being from a more deprived area, being part of a multiple birth, having a lower BW and not being breastfed all increased the likelihood of being left-handed. Further, being born in the summer/winter months and being born in England (as opposed to Scotland or Wales) also increased the likelihood of being left-handed. (Figure 2).

**Figure 2:**
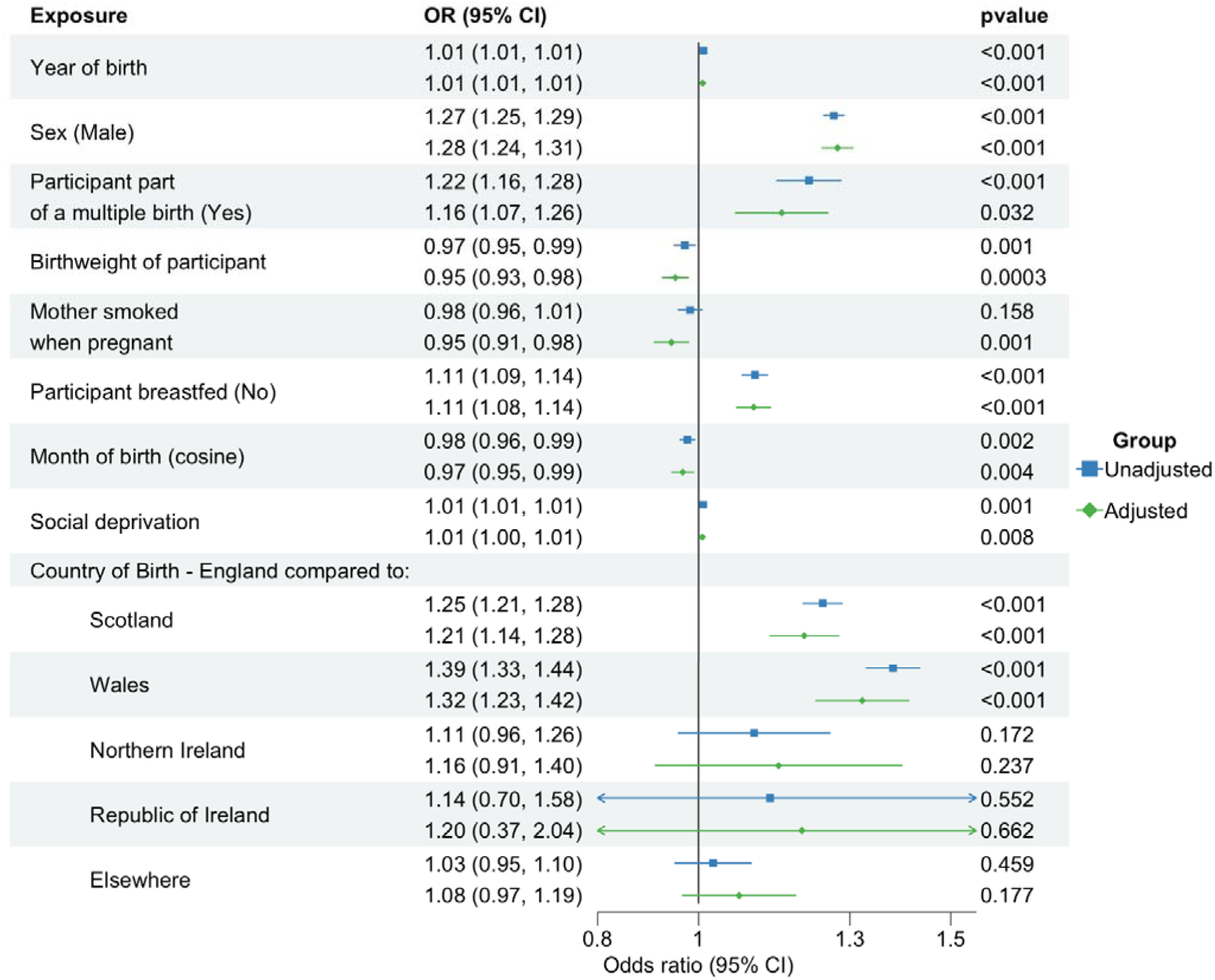
Association between early-life predictors of handedness and left-hand preference. An odds ratio > 1 indicates propensity to be left-handed. Unadjusted refers to results from univariable logistic regression analyses. Adjusted refers to results from multivariable logistic regression analyses.

Results from the multivariable analysis where all predictor variables and the first ten genome-wide principal components were included in the model, were similar to results from the univariable analysis. However, having a mother who smoked during pregnancy increased the likelihood of being right-handed after accounting for covariates. Results when stratifying by sex were similar to the whole cohort analysis and are presented in Figures 3 and 4 of the Supplement. The McFaddon pseudo R^2^ (*p*=0.005) suggested that the predictive power of the multivariable model, for individual hand preference was low. Results from the univariable and multivariable regression analyses are summarized in Figure 2.

**Figure 3:**
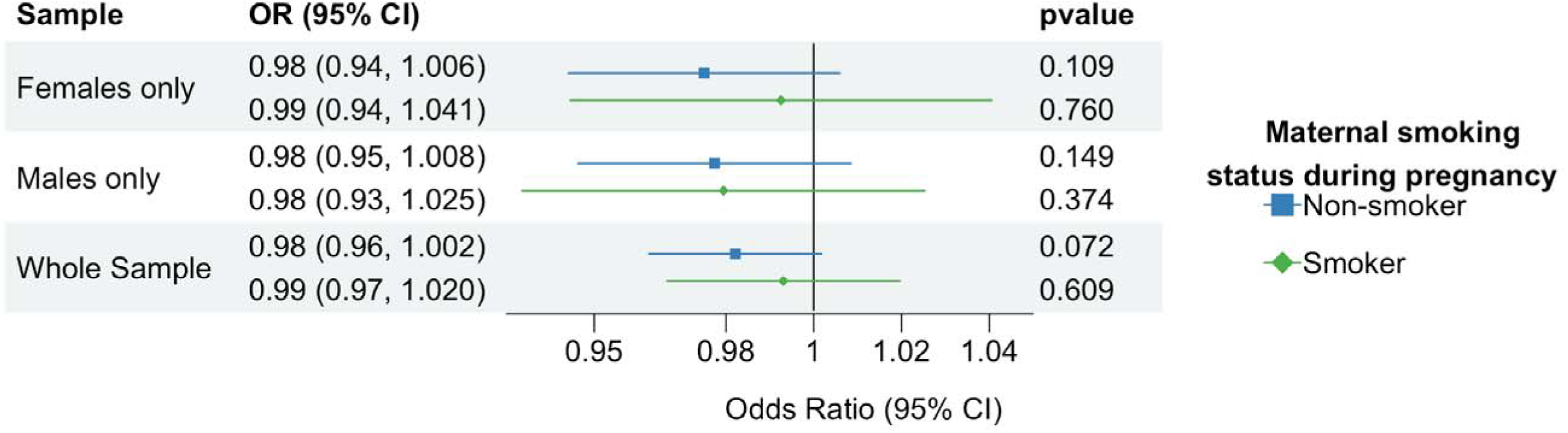
Odds ratio for offspring being left-handed per A allele at rs16969968. *Nb. Genetic association results show decreased odds of offspring being left-handed per additional allele A at rs16969968, adjusted for year of birth, sex and the first 10 principal components (N=346,943)*.

### Genetic Association Between Smoking Variants and Handedness

Demographic characteristics after exclusions are presented in Table 2. Characteristics stratified by mother’s smoking status are shown in Supplementary Table 5. Each additional smoking-increasing allele (additional allele A) at the rs16969968 locus increased the likelihood of maternal smoking during pregnancy (OR: 1.02; 95% CI: 1.004, 1.03; p-value: 0.0003). As a positive control and to demonstrate proof of principle for the GxE approach, we regressed birthweight on the SNP rs16969968. This is because, it is generally accepted that maternal smoking in pregnancy leads to lower offspring birthweight [34]. Therefore, if the SNP is a true proxy for maternal smoking behaviour during pregnancy, we should see a negative association with offspring birthweight in offspring of mothers who smoked during pregnancy. As expected, we found each additional smoking-increasing allele was associated with lower offspring birthweight in mothers who were reported to have smoked during pregnancy (beta: -0.01 kg per cigarettes per day; 95% CI: -0.02, -0.005; p-value: 0.001). However, there was evidence of an association between the SNP and being part of a multiple birth when conditioning on maternal smoking during pregnancy (Supplementary Table 6). This is likely to be an effect of collider bias and is examined in greater detail in the discussion.

In the unadjusted and models adjusted for year of birth, sex and the first ten principal components, we found no strong association between rs16969968 and offspring hand preference in the offspring of mothers who smoked during pregnancy or in the offspring of mothers who did not smoke during pregnancy. Sex stratified results were similar to the whole cohort analysis. Results from model 1 (unadjusted) are presented in Table 5 and model 2 (adjusted for year of birth, sex and the first ten genetic principal components) are presented in Figure 3.

**Table 5:** Genetic association between rs16969968 and left handedness in the unadjusted model.

|  |  | **OR** | **95% CI** | ***p*** |
| --- | --- | --- | --- | --- |
| Whole Sample | Non-smokers | 0.98 | 0.96, 1.002 | 0.083 |
|  | Smokers | 0.99 | 0.97, 1.02 | 0.682 |
| Females only | Non-smokers | 0.98 | 0.94, 1.01 | 0.11 |
|  | Smokers | 0.99 | 0.95, 1.04 | 0.818 |
| Males only | Non-smokers | 0.98 | 0.95, 1.01 | 0.167 |
|  | Smokers | 0.98 | 0.94, 1.03 | 0.459 |

*Nb: The odds ratio refers to being left-handed and the effect of the “A” allele at rs16969968.*

We then estimated the causal effect of maternal smoking during pregnancy on offspring hand preference using the Wald Ratio. We estimated the causal estimate in offspring who reported that their mothers smoked during pregnancy, as well as offspring who reported their mothers did not smoke during pregnancy. Under certain assumptions (e.g., the absence of collider bias, that maternal smoking does not affect offspring handedness pre-conception and postnatally etc) the Wald ratio in this latter group should provide an estimate of bias due to pleiotropy, since any effect of the SNP on hand preference in offspring cannot be due to maternal smoking behaviour during pregnancy. We therefore calculated the estimated causal effect of maternal smoking during pregnancy on offspring handedness accounting for pleiotropy by subtracting the estimated causal effects in the offspring of mothers who did and did not smoke during pregnancy. Again, we fit two models: model 1 which provided unadjusted estimates and model two, where the SNP-exposure and SNP-outcome associations were adjusted for sex, year of birth and the first ten principal components. Results are presented in Table 6.

**Table 6:** Estimated causal effect of maternal smoking heaviness during pregnancy on offspring left-hand preference.

|  | Causal effect of genetically predicted smoking heaviness on offspring left-hand preference in mothers who smoked during pregnancy |  | Causal effect of genetically predicted smoking heaviness on offspring left-hand preference in mothers who did not smoke during pregnancy |  | Causal effect of genetically predicted smoking heaviness on left-hand preference accounting for pleiotropy |  |
| --- | --- | --- | --- | --- | --- | --- |
| | $\beta_{IV}$ | 95% CI | $\beta_{IV}$ | 95% CI | $\beta_{IV}$ | 95% CI |
| rs16969968 model 1 | -0.01 | -0.06, 0.04 | -0.03 | -0.07, 0.005 | 0.02 | -0.04, 0.09 |
| rs16969968 model 2 | -0.01 | -0.07, 0.04 | -0.04 | -0.07, 0.003 | 0.02 | -0.04, 0.09 |
| PRS model 1 | -0.01 | -0.06, 0.04 | -0.02 | -0.06, 0.01 | 0.01 | -0.05, 0.07 |
| PRS model 2 | -0.01 | -0.06, 0.04 | -0.02 | -0.06, 0.01 | 0.01 | -0.05, 0.07 |

NB: Results show the log odds of offspring left handedness per cigarette smoked per day during pregnancy in mothers as calculated using the Wald estimator. Model 1 refers to the Wald estimator calculated without adjustment for covariates. Model 2 refers to the Wald estimator adjusting for sex, year of birth and the first 10 principal components (in the regression coefficients in both the numerator and denominator).

The PRS was slightly more predictive of own cigarettes smoked per day among self-reported smokers as reported in the UKB, compared to the single SNP rs16969968 (R^2^=0.005 vs 0.004). As in the single SNP analyses, we found a positive association between the PRS and maternal smoking during pregnancy and a negative association between the PRS and birthweight, our positive control (Supplementary Table 6). We did however, also find associations between the PRS and phenotypes not plausibly caused by smoking, such as social deprivation (Supplementary Table 6). Such implausible associations have been reported previously [35] and may reflect upstream effects captured in the original smoking GWAS and/or horizontal pleiotropy. A positive association between the smoking heaviness variants and year of birth appeared when stratifying by maternal smoking status. However, this is likely to be a collider effect (see Discussion).

We found no strong association between the PRS and offspring handedness in mothers who smoked during pregnancy or in mothers who did not smoke during pregnancy (Table 7a). We found the PRS was negatively associated with birthweight, and this was only in mothers that smoked during pregnancy (Table 7b). As in the single-SNP analysis, we used the Wald Ratio to estimate the causal effect of maternal smoking during pregnancy on offspring hand preference and to estimate the causal estimate in mothers that did not smoke during pregnancy (the pleiotropic effect). We were then able to estimate the causal effect of the maternal smoking during pregnancy on offspring hand preference, accounting for pleiotropy. We found there was no strong causal effect of maternal smoking during pregnancy on offspring hand preference in either subgroup. Results are presented in Table 6.

**Table 7a:**
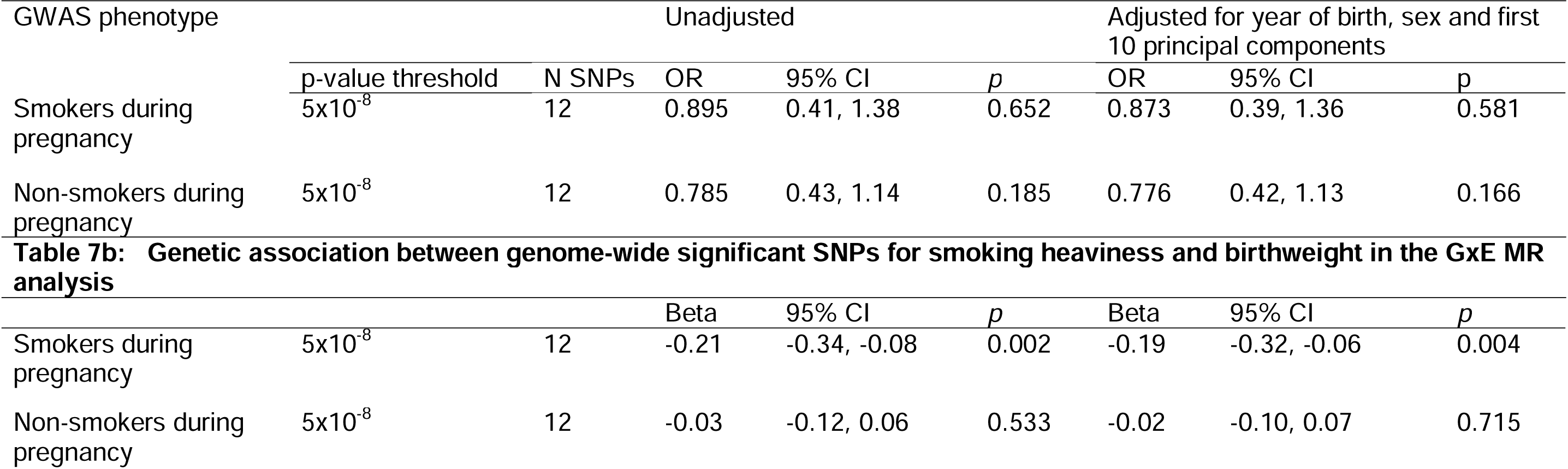
Genetic association between the PRS for smoking heaviness and left handedness.

## DISCUSSION

In this study, we investigated the relationship between early life exposures and handedness with a specific focus on maternal smoking during pregnancy. Our logistic regression analyses suggested that the odds of being left-handed were higher in males, those that were part of a multiple birth, had a lower birthweight or were bottle fed rather than breastfed. Additionally, social deprivation, being born more recently, being born in the summer/winter months or being born in England (as opposed to those born elsewhere) also increased the likelihood of being left-handed. These findings were maintained after controlling for the other predictors of handedness in the multivariable logistic regression analyses and are consistent with previous investigations in the UK Biobank [25]. However, we also found evidence to suggest that, after adjusting for covariates, and contrary to expectation, maternal smoking during pregnancy was associated with an increased likelihood of offspring being right-handed.

The unexpected result that maternal smoking during pregnancy is associated with an increased likelihood of offspring being right-handed, may reflect collider bias. Specifically, maternal smoking during pregnancy is associated with some of the variables we conditioned on in the multivariable model and is likely to be causal for some of these too e.g., birthweight [36]. If one or more unmeasured confounders were also causal for e.g., both birthweight and hand preference, then conditioning on birthweight in the multivariable model could generate a spurious association between maternal smoking during pregnancy and offspring hand preference. Indeed, when introducing birthweight into the multivariable model, the association between maternal smoking during pregnancy and offspring hand-preference became stronger which is consistent with this hypothesis.

We also investigated the association between maternal smoking during pregnancy and offspring handedness using genetic association analyses and a proxy GxE MR design [31]. These analyses suggested no strong causal effect of maternal smoking on offspring handedness, despite similar analyses showing a causal effect of maternal smoking on offspring birthweight (i.e. our positive control analysis). However, statistical power is limited in these designs for a number of reasons, complicating the interpretation of null results.

First, genetic variants typically explain only a small proportion of the phenotypic variance in the exposure variable, and consequently most MR studies require extremely large sample sizes to detect modest causal effects [37]. Further, proxy GxE MR studies use offspring variants as a surrogate for maternal genotype. As offspring variants are only expected to explain ∼25% of the variance in the maternal exposure compared to the same genotypes in the mother, power is reduced even further in this type of design. Third, proxy GxE MR studies require some sort of sample stratification for informative causal inference (i.e. maternal smoking status during pregnancy in the present study). This further reduces sample size and statistical power to detect a causal effect (particularly as mothers who smoked during pregnancy are a minority in the UK Biobank). Whilst it would be more statistically powerful to examine the relationship between directly genotyped mothers and their phenotyped offspring (simultaneously controlling for offspring genotype), this is only practically feasible in the very few cohorts that contain very large numbers of genotyped mother-offspring pairs [38–41]. For now, the proxy MR GxE design remains a valuable and practical addition to the expanding range of epidemiological and statistical methods aimed at enhancing the power to identify causal effects of maternal exposures on offspring outcomes [42].

One strategy we employed to increase the power of our MR analyses was to use a smoking-related PRS that explained more of the variance in maternal smoking heaviness. Although it is highly likely that at least some of the SNPs that comprise this score were pleiotropic (see Supplementary Tables 2 and 6 and [35]), we reasoned that an unbiased estimate of the causal effect might still be obtained by subtracting the estimated causal effect obtained in individuals whose mothers did not smoke during pregnancy from the causal effect obtained in the offspring of mothers who did smoke in pregnancy. However, one potential complication is that this strategy assumes that the variants used to proxy the exposure (smoking heaviness) are unrelated to smoking initiation or cessation (ever/never smoked). Our analyses showed that both the rs16969968 variant and the smoking heaviness PRS were both associated with the likelihood of mothers’ smoking during pregnancy-implying a relationship between the variants and ability to quit smoking. Additionally, some of the genetic variants have also been previously associated with smoking initiation (Supplementary Table 2). As we illustrate in Supplementary Figure 5, associations between genetic variants and smoking initiation/termination could result in collider bias when conditioning/stratifying on maternal smoking status during pregnancy. “Correcting” the causal estimates for pleiotropy (by the procedure outlined above) will not obviate this problem. The corollary is that our MR estimates are likely to have been biased through a collider effect.

There are some further limitations to our study. First, although the UKB is a prospective cohort, most of our variables were based on retrospective recall and self-report. Therefore, individuals may have been misclassified: e.g. participants may misreport if their mother smoked during pregnancy, especially due to the negative connotations surrounding such behaviour. We would expect that such misclassification might attenuate any association between maternal smoking and offspring handedness. Also, any misclassification will make the MR results from each group in the stratified analyses more similar and consequently reduce the effectiveness of any correction for pleiotropy.

Second, our analyses assume no relationship between the paternal genotype at rs16969968/GRS and offspring hand preference. However, any effect of paternal smoking on the foetus is likely to be far weaker than any maternal effect. Given that we found no effect of maternal smoking behaviour on offspring hand preference, we do not think that unmodelled paternal effects are a likely cause for concern.

Third we assume no assortative mating on smoking behaviour. It is well known that married couples tend to have more similar smoking habits than would be expected by chance [43]. Assortative mating induces complex patterns of correlations between smoking related loci between and within individuals [44]. The consequence is that phenotypic assortment can result in a myriad of possible paths between an ostensible instrumental variable and the outcome. Within family MR can generate causal estimates that are robust to assortative mating [45–47]. However, the GxE proxy MR design is not and as such, parental genotypes and behaviours may not be fully independent. Future work using mother-father-offspring genotype data could be used to explicitly model the parental genotype correlations and further refine the analysis. However, only in certain large cohorts is this possible [38, 39] and we believe our approach (removing related individuals and correcting for population stratification) adequately reduces the potential bias.

In conclusion, this study contributes to the understanding of handedness and its potential early-life determinants. Our findings using the UK Biobank cohort align with previous findings and emphasise the impact of factors such as birth year, birthweight, being part of a multiple birth and breastfeeding on hand preference. The results using the cohort data diverged from previous findings in that they suggested that maternal smoking during pregnancy may be associated with an increased likelihood of right-handedness rather than left-handedness. This unexpected finding, however, needs to be interpreted cautiously, considering the potential influence of collider bias. Additionally, when further investigating this association using a proxy GxE MR design, we found no strong evidence for a causal link between maternal smoking and handedness. However, it is unclear whether this is a true null effect or a consequence of limited power. Despite these challenges, our study highlights the complexity of the relationship between early-life exposures and handedness and emphasises the need for further exploration.

## Methods

### Study Participants

The UKB is a community-based, prospective study (https://www.ukbiobank.ac.uk) [19]. Recruitment of ∼500,000 participants and baseline assessments were completed between 2006-2010. Participants were aged 40-70 years at baseline, registered with a general practitioner and lived close to 22 assessment centres in England, Scotland, and Wales. Baseline assessments included demographics, lifestyle, and disease history, with linkages to electronic medical records. UK Biobank’s ethical approval was from the Northwest Multi-centre Research Ethics Committee.

We excluded individuals of non-white European ancestry (n=59,910) and those who had withdrawn their consent (n=52). We also excluded participants who reported a birthweight heavier than 6.0 kg (n=236) and lighter than 1 kg (n=1013) as, although some reports may have been accurate, the others would likely be self-report errors. This cut-off aimed to reduce the effects of outliers and measurement error in the model fitting. Participants were asked “Are you right or left-handed?” during a baseline questionnaire and self-reported their handedness (right-handed = 0, left-handed = 1 or ambidextrous = 2). Individuals who did not report their hand preference (n=92) were removed, as were individuals who reported being “ambidextrous” (n=7105) because reports of ambidexterity among participants are inconsistent across time points [25], and it is questionable the degree to which this simple self-report measure in UK Biobank reflects true ambidexterity (i.e. the ability to perform tasks equally well with either hand). We therefore had 434,104 individuals available for our study (see Figure 1 of the Supplement for flowchart of exclusions).

### Traditional Observational Epidemiological Analyses

#### Potential early life predictors of handedness

Exposure variables examined for being potential early life predictors of handedness were as follows: sex, being part of a multiple birth, maternal smoking while pregnant, if the participant was breastfed, country of origin (England, Wales, Scotland, Republic of Ireland, Northern Ireland or Elsewhere), year of birth, birth weight, month of birth and social deprivation. We modelled the month of birth as a cosine function in order to represent a continuous seasonal effect (i.e. months next to each other are more similar than other months) with peaks in the UK summer and winter (see [25] for more details). Social deprivation was modelled using the Townsend score (area-level deprivation) and was derived from data on unemployment, car ownership, household overcrowding and owner occupation aggregated at postcode area [48]. Townsend scores were assigned to participants based on their address at recruitment and were calculated immediately prior to recruitment using data from the preceding national census data (2001). Higher Townsend scores equate to higher levels of socioeconomic deprivation. All the variables were self-reported (other than sex and date of birth which were acquired from the central registry at recruitment). Responses of “do not know” and “prefer not to answer” were treated as missing values. See Supplementary Table 1 for the variables included in the analysis and how each variable was coded.

#### Statistical analysis

We performed univariable logistic regressions between each of the variables and hand preference. We then removed all participants with missing values for any of the predictor variables (final sample: n=193,770, 37.93 % male). The skew in the sex ratio is a result of males under reporting their birthweight (20.18% increase in missingness). Additionally, we performed a multivariable analysis of hand preference, where all variables were included in the model, adjusted for the first ten genome-wide principal components based on the genome-wide SNP data. We ran the multivariable analyses on the whole sample and in sex stratified samples. We also computed the McFadden pseudo R^2^ to assess how well the adjusted model explains the variation in the outcome.

Finally, we investigated the pairwise relationships between predictor variables and hand preference. We used Cramer’s V for categorical pairs, Pearson’s correlation coefficient for continuous variables, and Spearman’s rho when a pair comprised one categorical variable and one continuous variable. When examining the correlation between country of birth and the other variables, we looked at each location individually.

### Proxy Gene by Environment Mendelian Randomization

#### Genotyping and quality control

DNA was extracted from stored blood samples that had been collected from participants on their first visit to a UKB assessment centre. Genotyping was carried out using the UK Biobank Axiom Array by Affymetrix Research Services Laboratory in 106 sequential batches of approximately 4,700 samples. This resulted in a set of genotype calls for 489,212 samples and approximately 850,000 variants were directly measured. The UKB performed imputation centrally using the Haplotype Reference Consortium [49], UK10K [50], and 1000 Genomes project reference panels [51]. Genotype data were screened for genotyping quality (described elsewhere [51]), Hardy-Weinberg equilibrium failure (p<5×10^-8^) and minor allele frequency (MAF <0.01). SNPs used in our analysis were high-quality HRC-imputed dosage data provided by the UKB full release (IMPUTE4 INFO score ≥ 0.976) and extracted using plink 2.0 [52].

Participants were excluded following the same criteria as in the observational univariable logistic regression analysis. We also removed samples that had sex chromosome aneuploidy, a mismatch between genetically inferred sex and self-reported sex, high genotype missingness and samples that were excluded from kinship inference and autosomal phasing. Further, we removed individuals who had extreme heterozygosity as this can be an indication of poor-quality genotyping, sample contamination etc. One individual from each pair of participants whose genetic relatedness was inferred to be 3rd degree or closer were also excluded. This was done based on genotype data at SNPs spanning the genome, as previously calculated by Bycroft *et al* [53]. This left a sample of 346,871 individuals (see Figure 2 of the Supplement for a flowchart of exclusions).

#### Smoking phenotypes

At baseline assessment, participants were asked the question ‘Did your mother smoke regularly around the time when you were born?’. This was used to index maternal smoking during pregnancy.

#### Outcomes in participants

Handedness of participants was reported at baseline. As in the traditional observational epidemiological analysis, due to inconsistencies in the reporting of ambidexterity, only individuals who reported being left- or right-handed were retained.

#### Single SNP genetic association and instrumental variable analyses

We used individuals’ genotype at rs16969968, which is robustly associated with smoking heaviness (measured as cigarettes per day) [30], as a proxy for maternal genotype at the same locus, and consequently heaviness of maternal smoking. The SNP was coded as the number of smoking heaviness-increasing alleles (allele A).

#### Polygenic risk score analyses

We used GWAS summary statistics for smoking heaviness generated from a meta-analysis of 60 genome wide association studies of European ancestry (UKB and 23andMe participants removed) [54]. Independent SNP signals were identified using PLINK (r^2^ = 0.001, clump-kb = 1000). We created a genome-wide significant weighted polygenic risk score (PRS) of smoking heaviness for each individual in our sample (UKB) and used this as a proxy for the mothers’ PRS scores. Individuals were scored on the total number of smoking increasing alleles they carried across all genome-wide significant clumped variants weighted by regression coefficients from the GWAS of smoking heaviness (i.e. a weighted PRS). See Supplementary Table 2 for the variants used to construct the PRS, their closest gene and putative function.

#### Statistical Analysis

First, we investigated whether the SNP rs16969968 was associated with the likelihood of the mother smoking during pregnancy using logistic regression. Second, we stratified the mothers by their smoking status during pregnancy and tested the association between rs16969968 and handedness in each group using univariable regression and multivariable regression adjusting for year of birth, sex and the first ten genetic principal components. We repeated this analysis stratified by sex. Given that the participant’s genotype should not affect hand preference through their own smoking behaviour, (i.e. assuming hand preference long precedes smoking initiation), we did not account for the participant’s own smoking status in analyses.

We replicated this analysis using our PRS of smoking heaviness. As in the single-SNP analysis, we first investigated whether the weighted PRS was associated with mother’s reported smoking status during pregnancy. We then stratified on mother’s reported smoking status during pregnancy and ran a univariable regression and a multivariable regression accounting for year of birth, sex and the first ten principal components.

We conducted follow-up single SNP analyses using *Regenie* [55] which can account for population structure and cryptic relatedness across participants. The methods and results for these analyses are presented in the Supplement and did not differ substantively from the main results.

As a positive control and to demonstrate proof of principle for this approach, we separately regressed birthweight on the SNP rs16969968 and the PRS of smoking heaviness. This is because it is generally accepted that maternal smoking in pregnancy leads to lower offspring birthweight [34]. Therefore, if the SNP/PRS is a true proxy for maternal smoking behaviour during pregnancy we should see a negative association with offspring birthweight in offspring of mothers who smoked during pregnancy. To investigate the exchangeability assumption, we regressed the other predictors of hand preference (sex, birth year, being part of a multiple birth, being breastfed, birth month, social deprivation, and country of origin) on rs16969968 and the PRS. Under the exchangeability assumption, these phenotypes should not be associated with the SNP or the PRS. In both the positive and negative control analyses, we included the first ten genome-wide genetic principal components because its plausible that the frequency of the variants and the rates of the phenotypes may vary across ancestries.

We estimated the causal effect of maternal smoking heaviness during pregnancy on offspring hand preference using the Wald Ratio. We did this separately for both rs16969968 and the PRS. For the denominator of the Wald ratio, we calculated the association between rs16969968 /PRS and own smoking heaviness (self-reported cigarettes per day) among smokers in the UK Biobank.

## Acknowledgments

This research has been conducted using data from UK Biobank.

## Data Availability

Data were obtained from the UK Biobank cohort, as part of research application 53641, with David Evans as the principal applicant. Summary and description of these data can be found at, http://biobank.ctsu.ox.ac.uk/crystal/. For the use of the data, application must be made to, http://www.ukbiobank.ac.uk/register-apply/.

