## supplement for "Does smoking during pregnancy influence offspring handedness? Extending gene-by-environment Mendelian randomisation to include polygenic risk scores"

**Proxy Gene x Environment MR analysis**

One of the problems with GxE analyses in general is reduced statistical power. Stratifying the sample e.g. by maternal smoking status as we have done here, means that the number of individuals available for analysis is markedly reduced. Removing related individuals further reduces power, therefore we also ran proxy GxE genetic association analyses with related individuals included as sensitivity analyses. To do this we used Regenie [2] which is software developed to analyse binary traits with unbalanced case-control ratios on a genome-wide scale whilst also modelling population structure and cryptic relatedness. Results were largely unchanged when including related individuals in the analysis, whereby there was no association between the SNP rs16969968 and offspring hand preference (OR:1.02; 95% CI: 0.99, 1.06; p-value: 0.625).

| **Supplementary Table 1: Variables included in analysis and coding information** | | | |
| --- | --- | --- | --- |
|  | UK Biobank code | Type | Additional information |
| Year of Birth | f.34.0.0 | Continuous | 1936-1971 |
| Birthweight | f.20022.0.0 | Continuous | 0.45-5.98 (kg) |
| Part of a multiple birth | f.1777.0.0 | Categorical | 0=No, 1=Yes |
| Breastfed | f.1677.0.0 | Categorical | 0=Yes, 1=No |
| Maternal smoking during pregnancy | f.1787.0.0 | Categorical | 0=No, 1=Yes |
| Country of Birth | f.1647.0.0 | Categorical | England treated as baseline (0=England), with all other countries coded by a series of dichotomous variables. |
| Sex | f.31.0.0 | Categorical | 0=Female, 1=Male |
| Month of birth | f.52.0.0 | Continuous | Modelling using a cosine function for the 12 months |
| Handedness |  | Categorical | 0=right-handed, 1= left-handed, 2=ambidextrous |

| **Supplementary Table 2: Variants used to construct the PRS, their closest gene and putative function** | | | | | | | | | | | |  |
| --- | --- | --- | --- | --- | --- | --- | --- | --- | --- | --- | --- | --- |
| **CHR** | **POS** | **RSID** | **EA** | **OA** | **EAF** | **Beta** | **SE** | **P** | **N** | **Closest Gene** | **Putative gene Function** | **Known association** |
| 8 | 27568560 | rs1565735 | A | T | 0.20 | 0.02 | 0.004 | 4.33x10^-8^ | 183196 | intergenic |  | Smoking cessation, smoking status, smoking initiation, cigarettes per day, height |
| 9 | 15812667 | rs10962170 | G | A | 0.43 | 0.02 | 0.003 | 1.71x10^-8^ | 183196 | *CCDC171* | DNA-binding transcription factor activity and obsolete signal transducer activity |  |
| 11 | 113448730 | rs4274224 | A | G | 0.55 | 0.02 | 0.003 | 2.68x10^-8^ | 183196 | *DRD2* | Synthesis regulation, storage, and release of dopamine |  |
| 11 | 46741267 | rs61884324 | A | G | 0.10 | 0.03 | 0.006 | 2.59x10^-9^ | 183196 | *F2, CKAP5* | Regulates microtubule chromosome attachments, encodes prothrombin protein | smoking initiation |
| 15 | 78559273 | rs2036527 | A | G | 0.37 | 0.10 | 0.003 | 2.1x10^-17^ | 183196 | *CHRNA5* | Affects response dopaminergic neurons during chronic nicotine exposure and withdrawal | Average tendency to become a heavy smoker, response to bronchodilator (medication that relieves the symptoms of asthma), pulmonary function measurement, forced expiratory volume, cotinine measurement |
| 16 | 52063640 | rs11076320 | C | A | 0.63 | 0.02 | 0.003 | 8.88x10^-10^ | 181256 | *LINC02911* | Intergenic non-protein coding |  |
| 16 | 89827742 | rs1108064 | G | C | 0.57 | 0.02 | 0.003 | 6.46x10^-10^ | 181256 | *SPIRE2* | Establishment of meiotic spindle localization |  |
| 17 | 29236995 | rs112178027 | T | C | 0.18 | 0.03 | 0.004 | 1.52x10^-8^ | 183196 | *CRYBA1, TWF1P1* | Protein homodimerization activity and structural constituent of eye lens | Platelet count, brain stem volume measurement |
| 19 | 40743318 | rs142449067 | C | T | 0.04 | 0.05 | 0.01 | 2.44x10^-8^ | 183073 | *C19ORFf54* |  |  |
| 20 | 32361562 | rs6141293 | C | T | 0.35 | 0.02 | 0.003 | 4.95x10^-12^ | 183196 | *ASXL1* | Involved in chromatin remodelling |  |
| 20 | 63355597 | rs2273500 | C | T | 0.15 | 0.04 | 0.005 | 9.06x10^-16^ | 176936 | *CHRNA4* | Involved in the synthesis of neuronal nictotinic acetylecholine receptor. | Cigarettes per day, nicotine dependence, parental longevity, chronic obstructive pulmonary disease |
| 21 | 39272184 | rs145104523 | T | C | 0.13 | 0.03 | 0.005 | 4.18x10^-8^ | 183196 | *BRWD1* | Epigenetic control of meiotic chromosome stability in females and haploid gene transcription during post meiotic sperm differentiation in males. | Intelligence, BMI, ADHD, ASD |
| Nb: SNPs presented here were used to construct the PRS. Beta refers to the increase in the number of cigarettes smoked per day per increaser allele, SE refers to standard error, EA refers to the effect allele, OA refers to the other allele (reference allele), p refers to the p-value, N describes the sample size across contributing cohorts. | | | | | | | | | | | | |

| **Supplementary Table 3: Distribution of observed characteristics at baseline assessment in UK Biobank for participants who have no missing phenotype values.** | | | |
| --- | --- | --- | --- |
| **Variable** | **Categories** | **N (mean for continuous variables and % for categorical variables)** | **Frequency of left-hand preference vs righthanded (%)** |
| **Sex** | Female | 119, 903 (62.08) | 8.90 |
|  | Male | 73252 (37.92) | 10.98 |
| **Part of a multiple birth** | No | 188,171 (97.42) | 9.64 |
|  | Yes | 4984 (2.58) | 11.50 |
| **Maternal smoking** | No | 136, 946 (70.90) | 9.80 |
|  | Yes | 56, 209 (29.10) | 9.42 |
| **Breastfed** | No | 58, 692 (30.39) | 10.35 |
|  | Yes | 134,463 (69.61) | 9.40 |
| **Birthweight** |  | 193,155 (3.33 kg) |  |
| **Birth Year** |  | 193,155 (1953) |  |
| **Birth Month** |  | 193,155 (June) |  |
| **Social deprivation index** |  | 193,155 (-1.64) |  |
| **Country of Origin** | England | 162,578 (84.17) | 10.04 |
|  | Wales | 9563 (4.95) | 7.03 |
|  | Scotland | 16333 (8.46) | 7.92 |
|  | Northern Ireland | 862 (0.44) | 8.35 |
|  | Republic of Ireland | 77 (0.04) | 7.79 |
|  | Elsewhere | 3742 (1.94) | 9.35 |

| **Supplementary Table 4**: Associations between predictor variables and handedness (right-handedness=0, left-handedness=1) | | | | | | | | | | | | | | |
| --- | --- | --- | --- | --- | --- | --- | --- | --- | --- | --- | --- | --- | --- | --- |
|  | Year of Birth | Birthweight | Part of a multiple birth  (0=No, 1=Yes) | Breastfed  (0=Yes, 1=No) | Maternal Smoking  (0=No, 1=Yes) | Month of birth (cosine) | Sex  (0=Female, 1=Male) | England | Wales | Scotland | Northern Ireland | Republic of Ireland | Elsewhere | Handedness |
| Year of Birth | 1 |  |  |  |  |  |  |  |  |  |  |  |  |  |
| Birthweight | 0.006* | 1 |  |  |  |  |  |  |  |  |  |  |  |  |
| Part of a multiple birth | -0.013** | -0.178** | 1 |  |  |  |  |  |  |  |  |  |  |  |
| Breastfed | 0.175** | -0.081** | 0.053 | 1 |  |  |  |  |  |  |  |  |  |  |
| Maternal smoking | 0.012** | -0.069** | 0.00002 | 0.090 | 1 |  |  |  |  |  |  |  |  |  |
| Month of birth (cosine) | 0.009 | -0.020** | -0.001 | 0.011** | 0.003 | 1 |  |  |  |  |  |  |  |  |
| Sex | 0.006* | 0.153** | 0.006* | 0.039** | 0.012** | 0.0002 | 1 |  |  |  |  |  |  |  |
| England | -0.006* | 0.009 ** | 0.003** | 0.030 | 0.020 | 0.001 | 0.022 | 1 |  |  |  |  |  |  |
| Wales | -0.025** | -0.002 | 0.002** | 0.032 | 0.003 | -0.004 | 0.002 | NA | 1 |  |  |  |  |  |
| Scotland | -0.028** | 0.006* | 0.002** | 0.061** | 0.027** | 0.001 | 0.009 | NA | NA | 1 |  |  |  |  |
| Northern Ireland | 0.0009 | 0.004 | 0.0003 | 0.022** | 0.001 | 0.001 | 0.0009 | NA | NA | NA | 1 |  |  |  |
| Republic of Ireland | -0.012** | 0.004 | 0.009** | 0.024** | 0.005* | -0.003 | 0.005 | NA | NA | NA | NA | 1 |  |  |
| Elsewhere | 0.065** | -0.023** | 0.009** | 0.060** | 0.066** | 0.005* | 0.025** | NA | NA | NA | NA | NA | 1 |  |
| Handedness | 0.017** | -0.003 | 0.010** | 0.017** | 0.004 | -0.006 | 0.034** | 0.036** | 0.019** | 0.019** | 0.002 | 0.004 | 0.020** | 1 |
| For associations between categorical variables, Cramer’s V is presented. Associations between continuous variables are shown as Pearson R. Associations between categorical and continuous variables are shown as Spearman rho.  * = P < 0.05  ** = P < 0.001 | | | | | | | | | | | | | | |

| **Supplementary Table 5: Distribution of observed characteristics at baseline assessment in UK Biobank participants** | | | | | |
| --- | --- | --- | --- | --- | --- |
|  |  | **Mother smoked during pregnancy** | | **Mother who did not smoke during pregnancy** | |
| **Variable** | **Categories** | **N (mean for continuous variables and % for categorical variables)** | **Frequency of left-hand preference (%)** | **N (mean for continuous variables and % for categorical variables)** | **Frequency of left-hand preference (%)** |
| Number of smoking heaviness increasing alleles | 0 | 62903 | 9.81 | 115,332 | 9.94 |
|  | 1 | 62206 | 9.81 | 112,837 | 9.94 |
|  | 2 | 15410 | 9.67 | 27,616 | 9.47 |
| Sex | Male | 73,763 | 11.03 | 116,784 | 9.94 |
|  | Female | 66,756 | 8.68 | 139,001 | 8.95 |
| Year of birth |  | 140,519 (1951.37) |  | 255,785  (1950.99) |  |
| Social deprivation |  | 140,359  (-1.29) |  | 140,359  (-1.65) |  |

| **Supplementary Table 6.1: Association between rs16969968 and weighted PRS for smoking heaviness and binary predictor variables** | | | | | | | | | |
| --- | --- | --- | --- | --- | --- | --- | --- | --- | --- |
|  |  | **Whole cohort** | | | | **Smokers during pregnancy** | | | |
| **Threshold** | **exposure** | **OR** | **95% CI** | ***p*** | **R^2^** | **OR** | **95% CI** | ***p*** | **R^2^** |
| rs16969968 | Maternal smoking during pregnancy | 1.02 | 1.01, 1.03 | 3.1x10^-4^ | 3.5x10^-5^ |  |  |  |  |
|  | Sex | 0.996 | 0.99, 1.01 | 0.410 | 1.4x10^-6^ | 0.99 | 0.97, 1.01 | 0.203 | 8.33x10^-6^ |
|  | Breastfed | 1.00 | 0.99, 1.01 | 0.945 | 1.5x10^-8^ | 0.997 | 0.98, 1.02 | 0.753 | 7.8x10^-7^ |
|  | Part of a multiple birth | 1.03 | 0.99, 1.06 | 0.115 | 3.5x10^-5^ | 1.07 | 1.01, 1.12 | 0.02 | 2.0x10^-3^ |
| 5x10^-8^ | Maternal smoking during pregnancy | 1.87 | 1.65, 2.08 | 1.37x10^-8^ | 7.13x10^-3^ |  |  |  |  |
|  | Sex | 0.79 | 0.60, 0.97 | 0.010 | 1.20x10^-4^ | 0.72 | 0.43, 1.01 | 0.026 | 1.66x10^-4^ |
|  | Breastfed | 0.96 | 0.73, 1.19 | 0.745 | 5.74x10^-3^ | 1.02 | 0.66, 1.39 | 0.898 | 5.12x10^-3^ |
|  | Part of a multiple birth | 2.51 | 1.88, 3.15 | 0.004 | 4.39x10^-4^ | 4.18 | 3.19, 5.16 | 0.005 | 7.2x10^-4^ |
| **Supplementary Table 6.2: Association between rs16969968 and PRS for smoking heaviness and continuous predictor variables** | | | | | | | | | |
|  | **exposure** | **beta** | **95% CI** | ***p*** | **R^2^** | **beta** | **95% CI** | ***p*** | **R^2^** |
| rs16969968 | Birthweight | -0.01 | -0.001, -0.004 | 0.009 | -3.2x10^-6^ | -0.01 | -0.02, -0.01 | 0.001 | 0.0001 |
|  | Year of Birth | 0.01 | -0.03, 0.05 | 0.576 | -2.0x10^-6^ | 0.07 | 0.01, 0.12 | 0.03 | 2.49x10^-5^ |
|  | Social deprivation | 0.01 | -0.01, 0.02 | 0.473 | -1.4x10^-6^ | -0.01 | -0.03, 0.02 | 0.479 | -3.56x10-6 |
|  | Birth month (cosine) | 0.001 | -0.002, 0.01 | 0.537 | -1.87x10^-6^ | -0.002 | -0.01, 0.003 | 0.461 | -3.24x10^-6^ |
| 5x10^-8^ | Birthweight | -0.112 | -0.19, -0.04 | 0.005 | 0.001 | -0.20 | -0.33, -0.07 | 0.003 | 0.001 |
|  | Year of Birth | 0.477 | -0.26, 1.21 | 0.202 | 0.003 | 1.60 | 0.49, 2.71 | 0.005 | 0.004 |
|  | Social deprivation | 0.28 | 0.01, 0.55 | 0.042 | 0.013 | 0.20 | -0.24, 0.64 | 0.368 | 0.012 |
|  | Birth month (cosine) | 0.052 | -0.01, 0.12 | 0.112 | 3.74x10^-5^ | -0.004 | -0.11, 0.10 | 0.933 | -1.76x10^-5^ |
| NB: All associations adjusted for the first ten genetic principal components. The difference in size of the OR/beta between the single SNP and the PRS is partly due to the PRS being weighted. | | | | | | | | | |

UKB Cohort (N=502,512)

Excluded individuals:

Withdrawn consent: 52

Individuals not of White European ancestry: 59,910

Individuals of White European ancestry only

(N=442,550)

Individuals with birthweight between 1kg and 6kg (441,301)

Excluded individuals:

Individuals that report ambidexterity: 7105

Individuals who did not report hand preference: 92

Excluded individuals:

Individuals with birthweight greater than 6kg: 236

Individuals with birthweight less than 1kg: 1013

Left-handed and right -handed individuals only (N=434,104)

Figure 1: Flow diagram of exclusion criteria for traditional observational analyses

UKB Cohort (N=502,512)

Excluded individuals:

Withdrawn consent: 52

Individuals not of White European ancestry: 59,910

Individuals of White European ancestry only

(N=442,550)

Individuals with birthweight between 1kg and 6kg (441,301)

Excluded individuals:

Individuals that report ambidexterity: 7105

Individuals who did not report hand preference: 92

Left-handed and right -handed individuals only (N=434,104)

Excluded individuals:

Individuals with birthweight greater than 6kg: 236

Individuals with birthweight less than 1kg: 1013

Left-handed and right -handed individuals with genetic QC (N=432,519)

Excluded individuals:

Related individuals: 81,369

No genetic data [1]: 4279

*Figure 2: Flow diagram of exclusion criteria for proxy GxE MR analyses*

Excluded individuals:

Aneuploidy: 631

Excluded from Kinship: 787

Heterozygosity outliers: 968

Sex mismatch: 166

(Some individuals fall into more than one category. Total individuals removed: 1493)

Unrelated left-handed and right -handed individuals with genetic QC (N=346,871)

**Figure 3**: Association between early-life predictors of handedness and left hand preference in females only. An odds ratio > 1 indicates propensity to be left-handed. Unadjusted refers to results from univariable logistic regression analyses. Adjusted refers to results from multivariable regression analyses.

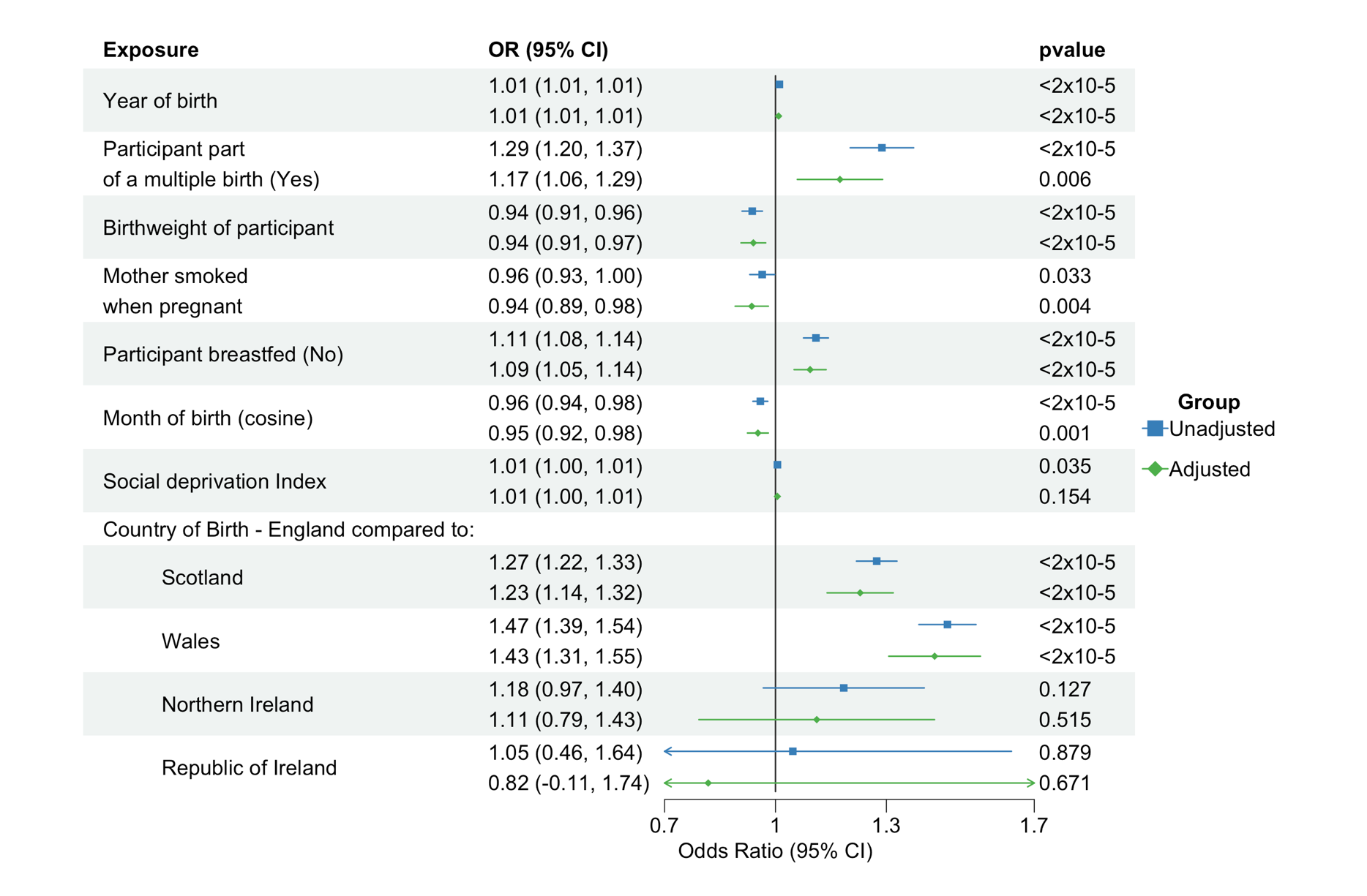

**Figure 4**: Association between early-life predictors of handedness and left hand preference in males only. An odds ratio > 1 indicates propensity to be left-handed. Unadjusted refers to results from univariable logistic regression analyses. Adjusted refers to results from multivariable regression analyses.

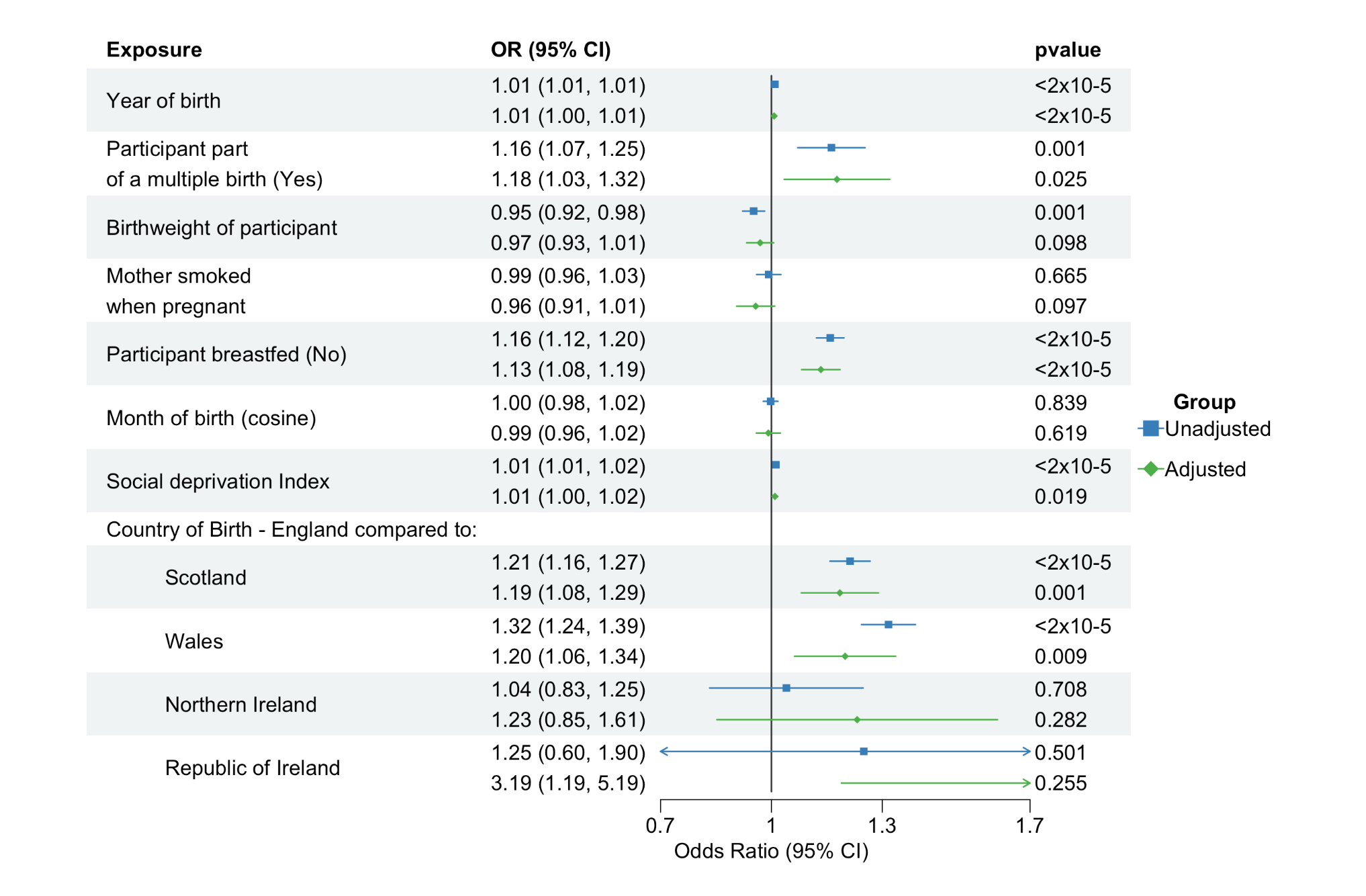

Figure 5: Directed Acyclic Graph to demonstrate potential collider bias in Gene by Environment Mendelian randomization analyses.

Diagram illustrating the potential issue of collider bias in the gene x environment Mendelian randomisation (GxE MR) analysis. The issue will arise if SNPs that are related to smoking initiation/termination are used as instruments in the analysis. In the GxE model, we condition on smoking status during pregnancy. If there are additional confounders (C) that cause both smoking initiation/termination (ever/never smoked) and offspring hand preference, this will create a pathway between the SNP and offspring hand preference when SNPs included in the model are associated with smoking initiation/termination as well as maternal smoking heaviness. This is because smoking initiation/termination becomes a collider variable.

SNP

C

Offspring hand preference

Maternal Smoking heaviness

Smoking initiation/termination

Unmeasured confounders

1.	Bycroft, C., et al., *The UK Biobank resource with deep phenotyping and genomic data.* Nature, 2018. **562**(7726): p. 203-209.

2.	Mbatchou, J., et al., *Computationally efficient whole-genome regression for quantitative and binary traits.* Nature Genetics, 2021. **53**(7): p. 1097-1103.
